# Proteogenomic analysis of 5,411 plasma proteins in sickle cell disease patients

**DOI:** 10.64898/2026.04.06.26350255

**Authors:** Cristian Groza, Arnaud Chignon, Ken Sin Lo, Vanessa Bellegarde, Pablo Bartolucci, Guillaume Lettre

**Affiliations:** Université de Montréal, 2900 Boul. Édouard-Montpetit, Montréal, Québec, H3T 1J4, Canada; Montreal Heart Institute, 5000 Bélanger Street, Montréal, Québec, H1T 1C8, Canada; Red Cell Genetic Disease Unit, Hôpital Henri-Mondor, Assistance Publique-Hôpitaux de Paris (AP-HP), Université Paris Est, IMRB - U955 - Équipe no 2, Créteil, France

**Keywords:** Sickle cell disease, proteome, pQTL, fetal hemoglobin

## Abstract

There are few therapeutic options to treat patients with sickle cell disease (SCD), a blood disorder caused by mutations in the β-globin gene that affects >7M individuals worldwide. Combining human genetics and high-throughput proteomics can help identify new drug targets. Here, we present results from a proteogenomic analysis of the plasma proteome in SCD patients. We measured the levels of 5,411 plasma proteins and tested their associations with common genetic variation in 343 SCD patients. After conditional analyses, we identified 560 protein quantitative trait loci (pQTL), including 58 (10%) that are novel. Many of these pQTL are not specific to SCD patients and associate with clinically relevant traits in non-SCD African Americans from the Million Veteran Program (e.g. hemoglobin concentration, triglycerides). The effect sizes of the pQTL is largely concordant between SCD and non-SCD individuals, although we found examples (e.g. APOL1, haptoglobin) with evidence of heterogeneity that suggests an interaction between the plasma proteome and the SCD genotype. Finally, we combine pQTL and genome-wide association study results for fetal hemoglobin (HbF) in a Mendelian randomization analysis to prioritize five proteins that may increase HbF production (ENPP5, LBP, NAAA, PT3X, ZP3).

## INTRODUCTION

Sickle cell disease (SCD) is the most common monogenic disease in the world, affecting >7M individuals, mostly in sub-Saharan Africa and the Indian subcontinent (GBD 2015 Disease and Injury Incidence and Prevalence Collaborators 2016). This blood disorder is caused by mutations in the β-globin gene and is characterized by extreme clinical heterogeneity, ranging from mild to life-threatening complications (e.g. overt stroke, acute chest syndrome) (Kato et al. 2018). Patients with SCD have a lower quality-of-life and life expectancy when compared to non-SCD individuals matched on ethnicity and socio-economic status (Lubeck et al. 2019).

Unfortunately, there are few therapeutic options to treat SCD patients. Curative interventions, including allogenic transplants and autologous gene therapy, remain complex and expensive, and are therefore not accessible to most SCD patients (Gaspar and Gaspar 2025). Blood transfusions (or apheresis) are effective to prevent major complications, but require regular visits to transfusion centers and are not without risks (e.g. allo-immunization, iron overload). Hydroxyurea, an inhibitor of ribonucleotide reductase, has emerged as a safe and effective drug that works in part by increasing the production of fetal hemoglobin (HbF) levels (Orah S. Platt 2008). HbF can replace sickled hemoglobin and has been shown to protect against most SCD-related complications, including painful crises, stroke and death (O. S. Platt et al. 1991, 1994; Sommet et al. 2016; Pincez and Lettre 2023). Not all SCD patients respond equally well to HU treatment (Steinberg et al. 1997; Ma et al. 2007). Two other drugs, the P-selectin inhibitor Crizanlizumab and the anti-oxidant metabolite L-glutamine, have been approved more recently to treat SCD patients, but are not yet widely used in practice. There is a clear need for additional affordable drugs to prevent the most severe SCD-related complications.

Proteogenomic approaches offer new opportunities to identify drug targets for human diseases (Suhre et al. 2021; Karim et al. 2026). It relies on the possibility to accurately measure 1000s of proteins in human biospecimens and correlate their levels with genetic variation. Already, efforts in mostly healthy populations have identified 1000s of protein quantitative trait loci (pQTL) for human plasma proteins (Tahir et al. 2024; Sun et al. 2023; Cai et al. 2025; Ferkingstad et al. 2021; Pietzner et al. 2021; Zhang et al. 2022). These pQTL discoveries are important for at least two reasons: First, when pQTL and disease genome-wide association study (GWAS) signals are co-localized, it suggests that genetic variation impacts disease risk through the measured proteins. Such a result can serve as a powerful strategy to prioritize GWAS causal genes and establish mechanisms of action. Second, we can use pQTL as instrument variables in Mendelian randomization (MR) analyses to infer causality. Positive MR results can distinguish between biomarkers and potential drug targets, and thus significantly help guide the development of new therapies.

In this study, we performed a proteogenomic experiment in patients with SCD. We identified novel pQTL and tested their associations with SCD-specific complications and with clinically-relevant traits measured in a non-SCD cohort (e.g. hematological and lipid traits). We confirmed that most pQTL found in non-anemic African-ancestry cohorts replicate in SCD patients. Finally, we used our pQTL discoveries and a meta-analysis of HbF GWAS results to prioritize plasma proteins that may modulate HbF production in SCD patients.

## RESULTS

### pQTL discovery in sickle cell disease patients

We measured 5,416 proteins in the plasma of 397 SCD patients of African ancestry from the GEN-MOD cohort using an antibody-based proteomic platform (Olink) (Chignon et al. 2026). After quality-control, we obtained data from 5,411 proteins and 343 SCD participants with genome-wide genotyping data available (**Methods**) (Ilboudo et al. 2017, 2025). All GEN-MOD participants had the HbSS genotype (**Table S1**). At recruitment, they were not treated with hydroxyurea nor had they received a blood transfusion in the last 90 days. We collected their plasma samples at steady-state. We tested the association between 4.6M imputed genetic variants and normalized protein levels using an additive model, correcting for age, sex and the first 20 principal components (**Methods**). We defined *cis*-pQTL as variants located <1-Mbp from the genes encoding the tested proteins, and used different thresholds to declare significance for *cis* and *trans* associations to account for multiple testing, consistent with a recent pQTL study (*P_cis_*<5×10^−8^, *P_trans_*<7.7×10^−11^) (Tahir et al. 2024). After conditional analyses (**Methods**), we identified 529 *cis*-pQTL and 31 *trans*-pQTL implicating 467 unique proteins (**Fig. 1A** and **Table S2**). Ten of the *trans*-pQTL implicated genetic variants at the highly pleiotropic *ABO* locus (**Table S2**) (Katz et al. 2022).

**Figure 1.**
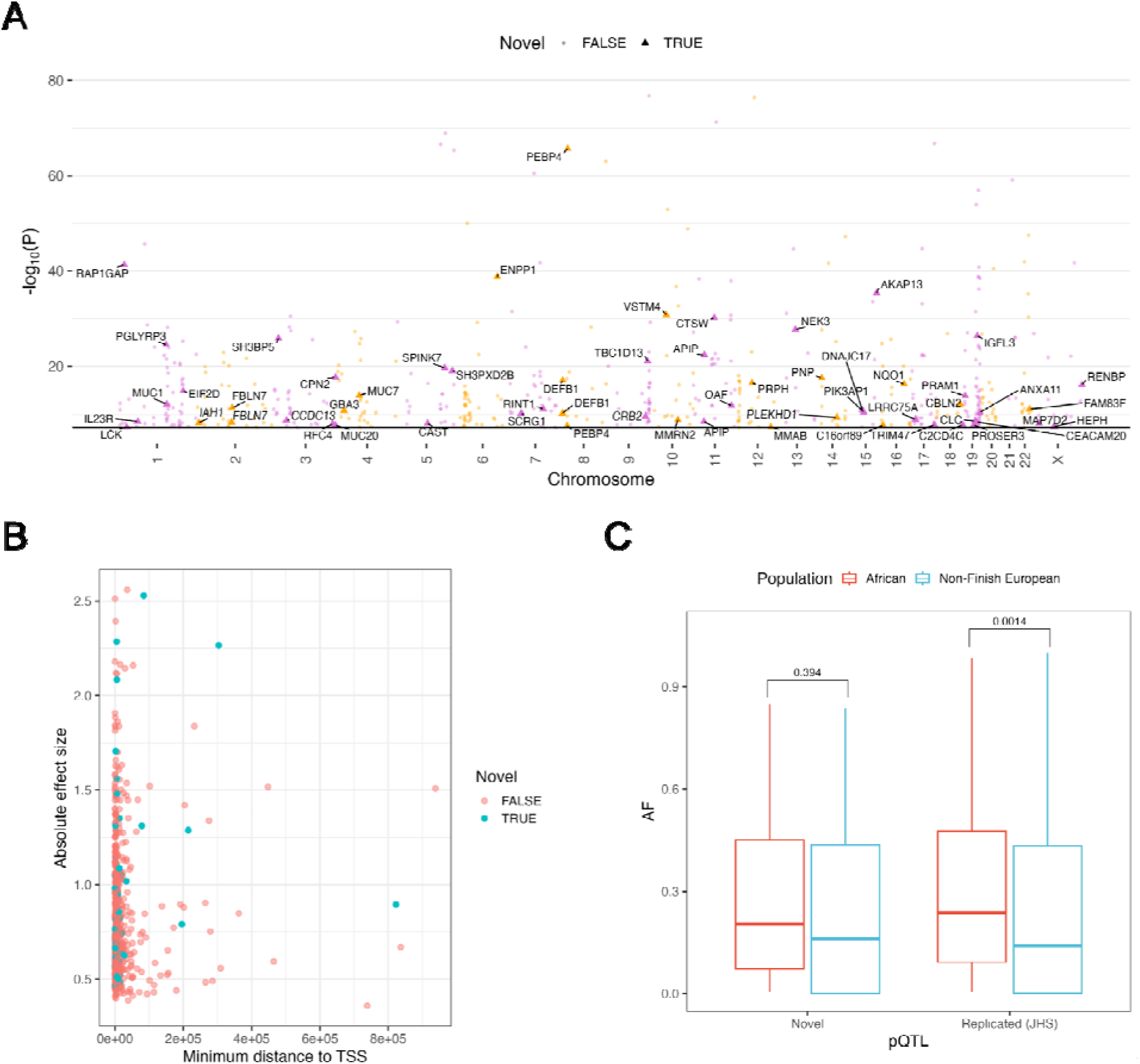
Identification and characterization of protein quantitative trait loci (pQTL) in sickle cell disease (SCD) patients from the GEN-MOD cohort. (**A**) Manhattan plot showing independent pQTL associated with protein levels in GEN-MOD. Highlighted are novel associations (triangles) and previously known associations (circles). We used different colors to distinguish between contiguous chromosomes. (**B**) Position of novel and known pQTL relative to the associated protein transcription start site (TSS). (**C**) Allele frequency (AF, gnomAD) of novel and known pQTL in African-ancestry individuals (AFR) contrasted with allele frequency in non-Finnish European ancestry individuals (NFE). We calculated statistical significance with Student’s *t*-test. The difference was not significant for the novel pQTL due to their small number (N=58 variants).

Tahir et al. reported pQTL GWAS results from the analyses of 2,881 plasma proteins measured with the Olink platform in 1,054 Black adults from the population-based Jackson Heart Study (JHS) (Tahir et al. 2024). We compared the pQTL results for the overlapping set of 2,721 proteins measured in both the JHS and our GEN-MOD SCD cohort. Of the 930 protein-variant pairs identified in the JHS and tested in GEN-MOD, we replicated 484 pQTL with consistent direction of effect and correction for multiple testing (binomial *P*<2.2×10^−16^)(**Table S3**). This replication rate in the smaller GEN-MOD cohort highlights the quality of our pQTL dataset.

### Novel pQTL identified in sickle cell disease patients

Of the 560 independent variant-protein pairs identified in GEN-MOD, 58 (10%) are novel (**Fig. 1A** and **Table S2**). For 51/58 (88%), the list of novel pQTL included proteins that were not tested on older Olink assays. The remaining seven pQTL involve association signals that are unreported and not in linkage disequilibrium (LD) with any pQTL variants in the JHS study or the large UK Biobank (Sun et al. 2023), although the proteins (ANXA11, CEACAM20, CLC, HEPH, PIK3AP1, RFC4, SCRG1) were measured in those cohorts (**Table S2**). This may indicate SCD-specific variant-protein associations, although replication is needed to address this possibility. Similar to the known variant-protein pairs identified in the JHS, the novel pQTL found in GEN-MOD tended to be located near the transcription start site of the genes encoding the tested proteins (**Fig. 1B**). Furthermore, the pQTL found in the African-ancestry GEN-MOD cohort implicated variants with minor allele frequencies more common in African- than in European-ancestry individuals from gnomAD (**Fig. 1C**).

We tested the association between the 560 pQTL found in GEN-MOD and five SCD-related complications (stroke, priapism, avascular necrosis, leg ulcers, cholecystectomy) in up to 1,641 SCD participants from GEN-MOD and the Cooperative Study of Sickle Cell Disease (CSSCD)(**Table S1**), but found no significant association (**Table S4**). To determine if the pQTL identified in GEN-MOD may be clinically important in non-SCD populations, we queried the corresponding variants in the phenome-wide association study (pheWAS) results from 121,177 African Americans that participate in the VA Million Veteran Program (MVP) (Verma et al. 2024). We found 58 (3 novel) pQTL variants from GEN-MOD that are associated with laboratory measures in MVP (**Table S5**). While many of these pQTL-pheWAS overlaps suggest new biology, several highlight plausible mechanisms: transferrin (TF) and red blood cell traits or angiopoietin-like-4 (ANGPTL4) and plasma lipid levels (**Table S5**). The overlap between a novel pQTL for Charcot-Leyden crystal (CLC) protein/Galectin-10 and a pheWAS signal for eosinophil counts is also interesting. CLC is one of the most abundant proteins within human eosinophils (Weller et al. 2020) and is involved in eosinophil granulogenesis (Grozdanovic et al. 2020). CLC is emerging as a new biomarker for disease activity, diagnosis and treatment effectiveness in asthma and several allergic reactions (Tomizawa et al. 2022).

### Comparison of effect sizes for pQTL in SCD and non-SCD individuals

The effect sizes of the pQTL were largely consistent between GEN-MOD and the JHS, suggesting that the HbSS genotype had limited impact on the associations with plasma protein levels (Pearson’s *r*=0.90, *P*<2.2×10^−16^)(**Fig. 2**). In fact, we only detected seven pQTL from the JHS study that had heterogeneous effect sizes when compared to GEN-MOD: four proteins with *cis*-pQTL (MSMB, APOL1, PTPRH, HSDL2) and three proteins with *trans*-pQTL (SERPIND1, PON3, GALNT2)(**Table 1**). For these variant-protein pairs, the difference in effect sizes could be due to the HbSS genotype, although it is also possible that differences in antibody affinity between the different Olink panels used for GEN-MOD and JHS may account for some of the observed heterogeneity.

**Figure 2.**
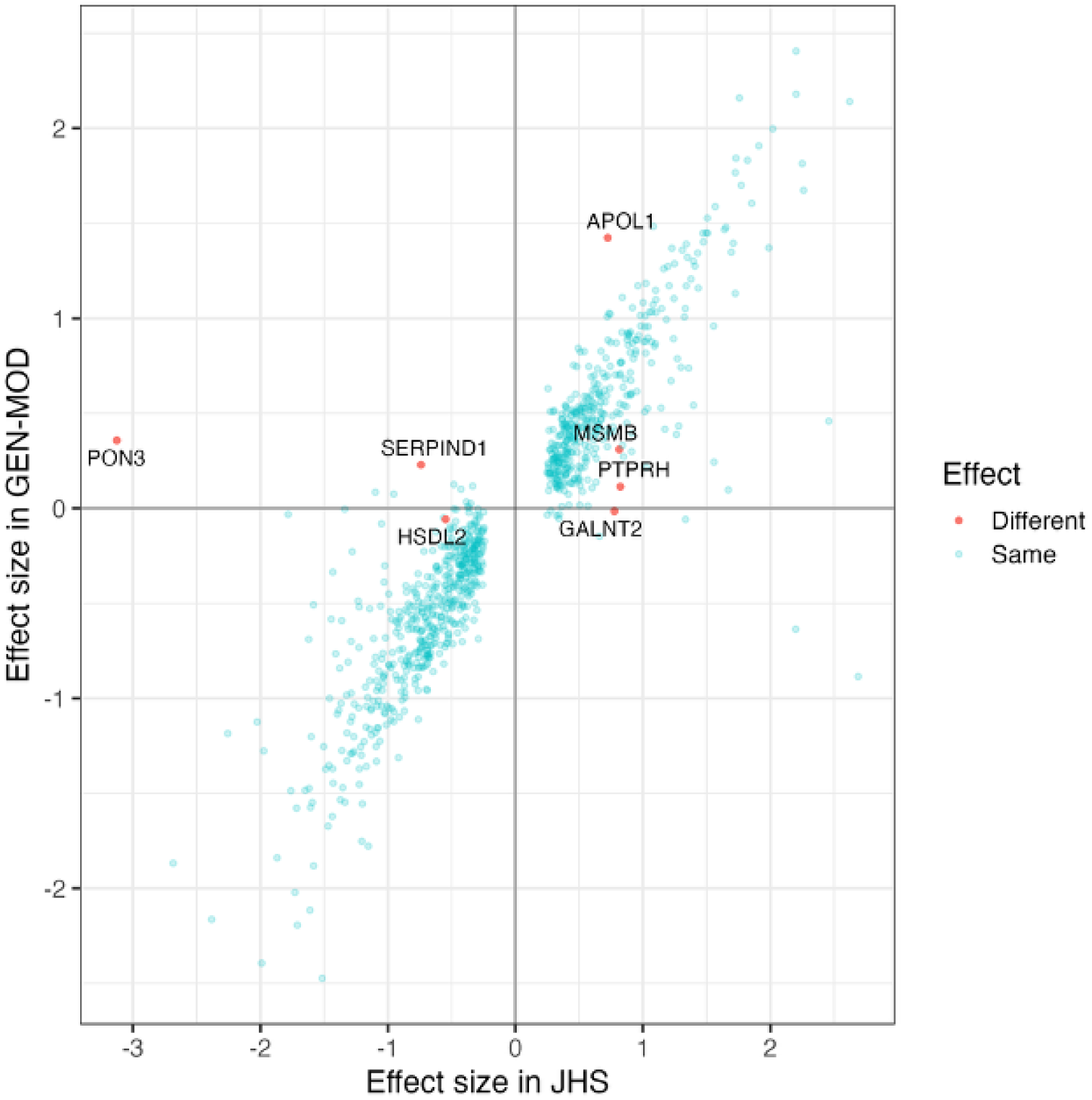
Protein quantitative trait locus (pQTL) effect sizes in non-anemic and sickle cell disease (SCD) populations. We compared the normalized effect size (per allele) of the genetic variants associated with plasma protein levels in the Jackson Heart Study (*x*-axis) and GENMOD (*y-*axis). Effect sizes are largely consistent, with only seven variant-protein pairs that show statistically significant heterogeneity (in red, with corresponding protein name).

**Table 1.** pQTL with heterogeneous effect sizes between the GEN-MOD sickle cell and the JHS population-based cohorts. All participants have genetically defined African ancestry. The effect allele is in bold in the VariantID column. Effect sizes (BETA) and standard errors (SE) are in standard deviation units.

| VariantID | Protein | pQTL | GEN-MOD |  | JHS |  | Het. P-value | Protein description |
| --- | --- | --- | --- | --- | --- | --- | --- | --- |
|  |  |  | BETA (SE) | P-value | BETA (SE) | P-value |  |  |
| chr9:112447393:C:**A** | HSDL2 | *cis* | -0.057 (0.082) | 0.49 | -0.548 (0.044) | 2.8x10<sup>-35</sup> | 1.5x10<sup>-7</sup> | Hydroxysteroid dehydrogenase-like protein 2, role in regulating lipid metabolism |
| chr10:46046326:A:**G** | MSMB | *cis* | 0.310 (0.077) | 6.6x10<sup>-5</sup> | 0.815 (0.046) | 7.2x10<sup>-70</sup> | 1.8x10<sup>-8</sup> | Member of the immunoglobulin binding factor family |
| chr19:55191542:C:**A** | PTPRH | *cis* | 0.114 (0.088) | 0.20 | 0.823 (0.047) | 2.6x10<sup>-67</sup> | 1.2x10<sup>-12</sup> | Protein tyrosine phosphatase receptor type H |
| chr22:36269923:C:**T** | APOL1 | *cis* | 1.424 (0.104) | 4.8x10<sup>-34</sup> | 0.724 (0.066) | 8.4x10<sup>-28</sup> | 1.3x10<sup>-8</sup> | Lipid exchange and transport throughout the body, trypanosome lytic factor |
| chr5:152294430:T:**C** | PON3 | *trans* | 0.358 (0.571) | 0.53 | -3.126 (0.417) | 6.6x10<sup>-14</sup> | 8.5x10<sup>-7</sup> | Paraoxonase-3, antioxidant enzyme |
| chr16:72054562:A:**C** | SERPIND1 | *trans* | 0.229 (0.138) | 0.098 | -0.740 (0.072) | 8.5x10<sup>-25</sup> | 4.6x10<sup>-10</sup> | Plasma serine protease, thrombin and chymotrypsin inhibitor, heparin cofactor 2 |
| chr16:72054562:A:**C** | GALNT2 | *trans* | -0.015 (0.140) | 0.92 | 0.776 (0.072) | 4.1x10<sup>-27</sup> | 5.0x10<sup>-7</sup> | Enzyme that catalyzes O-linked oligosaccharide biosynthesis |

The *trans*-pQTL for SERPIND1 and GALNT2 implicate the same variant, chr16:72054562:A:C, which is located just upstream of the haptoglobin (*HP*) gene on chromosome 16. HP is the plasma protein responsible for scavenging cell-free hemoglobin levels (Smith and McCulloh 2015), and its levels are low in SCD because of its depletion due to constant hemolysis (Santiago et al. 2018). It is not possible to determine directly in GEN-MOD or the JHS if chr16:72054562:A:C is a *cis*-pQTL for HP because the protein is not measured on the Olink platform. However, chr16:72054562:A:C is in linkage disequilibrium (LD, *r*^2^=0.56 in African Americans [ASW] from the 1000 Genomes Project) with rs77303550, a variant associated in *cis* with HP plasma levels as measured by mass spectrometry (Johansson et al. 2013) and in *trans* with SERPIND1 (Emilsson et al. 2018; Loya et al. 2025). rs77303550 has been associated with several lipid and hematological traits (Karjalainen et al. 2024; Loya et al. 2025; Jee et al. 2025). Based on these results, it is likely that chr16:72054562:A:C is a *cis*-pQTL for *HP* in healthy individuals, but is not associated with HP (nor SERPIND1 and GALNT2) in GEN-MOD because of the direct effect of SCD-related hemolysis and cell-free hemoglobin on HP levels (see **Discussion**).

Of the seven proteins with heterogeneous effect sizes, only the association with APOL1 levels is significant in SCD patients, with an effect size in GEN-MOD that is nearly double the effect size in the JHS (**Table 1**). In individuals of African descent, genetic variants in the apolipoprotein L1 (*APOL1*) gene – including the G1 and G2 coding alleles – have been implicated in resistance to trypanosomiasis but also increased risk of non-diabetic chronic and end-stage kidney disease (Limou et al. 2014; Genovese et al. 2010). In SCD patients, the G1/G2 *APOL1* genetic variants are associated with albuminuria and SCD nephropathy (Zahr et al. 2019; Ashley-Koch et al. 2011; Saraf et al. 2017). The sentinel pQTL variant for *APOL1* reported in the JHS (chr22:36269923:C:T, rs6000222) is in strong LD with the G2 allele (chr22:36265995:AATAATT:A / rs71785313) in GEN-MOD (*r*^2^=0.91), and both variants are associated with increased APOL1 plasma protein levels (*P*_rs6000222_=4.8×10^−34^ and *P*_rs71785313-G2_=6.2×10^−36^). The *APOL1* G1 allele (composed of two variants, chr22:36265860:A:G [rs73885319] and chr22:36265988:T:G [rs60910145]) is not associated with APOL1 protein levels in GEN-MOD (*P*>0.05). Given the strength of the pQTL association between rs6000222/rs71785313-G2 and APOL1 protein levels, we expected that the same variants would also be associated with estimated glomerular filtration rate (eGFR), a measure of kidney function. However, rs6000222, rs71785313-G2 and rs73885319/rs60910145-G1 are not associated with eGFR in GEN-MOD (*P*>0.05 for the additive and recessive genetic models), presumably because of the limited sample size (N_eGFR_=395). This suggests that the strong pQTL association between the G2 coding allele and APOL1 plasma levels as measured by Olink most likely reflects differential epitope:antibody binding rather than true changes in APOL1 protein levels. The impact of protein-altering variants on protein measurements done using affinity reagents (e.g. antibodies, aptamers) has been noted before (Nicholas et al. 2026).

### Association between pQTL and fetal hemoglobin levels

Fetal hemoglobin (HbF) is one of the main modifiers of disease severity in SCD (Lettre and Bauer 2016), and increasing HbF levels with hydroxyurea treatment is a cornerstone of preventive medicine in SCD. Finding additional drug targets to modulate HbF levels remains a top priority because not all patients respond equally well to hydroxyurea (Orah S. Platt 2008) and CRISPR-based gene therapy to disrupt the HbF regulator *BCL11A* is costly (Frangoul et al. 2024). Since plasma proteins are easy to measure and modulate, they would represent an interesting new class of molecules to investigate for a potential role in controlling HbF production in SCD.

We tested the association between baseline HbF levels and the levels of all 5,411 plasma proteins measured by Olink in 376 HbSS GEN-MOD participants. We found 14 proteins associated with decreased and one protein (CEACAM16) associated with increased HbF levels (false discovery rate [FDR]<5%)(**Table S6**). Of these 15 proteins, seven (ICAM4, IL1RL1, NRP2, OMD, CTH, MRI1, C7) were not associated with hemoglobin concentration, suggesting a more specific association with HbF levels rather than with overall hemoglobin production (**Table S6**). Literature review of the functions of these seven proteins did not yield obvious connections with HbF regulation nor erythropoiesis.

Next, we integrated the pQTL results with our recent meta-analysis of HbF GWAS results carried out in SCD patients (Ilboudo et al. 2025). For all downstream analyses, we excluded GEN-MOD from the HbF meta-analysis to ensure that pQTL and HbF genetic association results are independent. There was no enrichment of association signals in the HbF meta-analysis for the genome-wide significant pQTL identified in GEN-MOD (**Fig. 3A** and **Table S7**). Mendelian randomization (MR) can help distinguish mere associations from causal interactions. We performed two-sample MR analyses by selecting as instrumental variables genetic variants with a conditional pQTL P-value<0.001 that are located <500-kb from the transcriptional start site (TSS) of the encoded protein (**Methods**). We found 14 proteins with putative causal associations, including five proteins associated with increased HbF levels (**Table 2** and **Table S8**). For instance, our MR results implicate PTX3, a marker of acute inflammation and tissue ischaemia that is at elevated levels in SCD patients (Nur et al. 2011; Elshazly et al. 2014) and ENPP5, an enzyme involved in nucleotide metabolism, in the increased production of HbF in SCD patients (**Fig. 3B-C**). We also noted that a *cis*-pQTL variant for ENPP1, chr6:131851228:A:C, is nominally associated with HbF levels (P-value=0.010)(**Fig. 3A** and **Table S7**). The remaining three proteins implicated by MR in increased HbF productions are ZP3, LBP and NAAA (**Table 2**).

**Figure 3.**
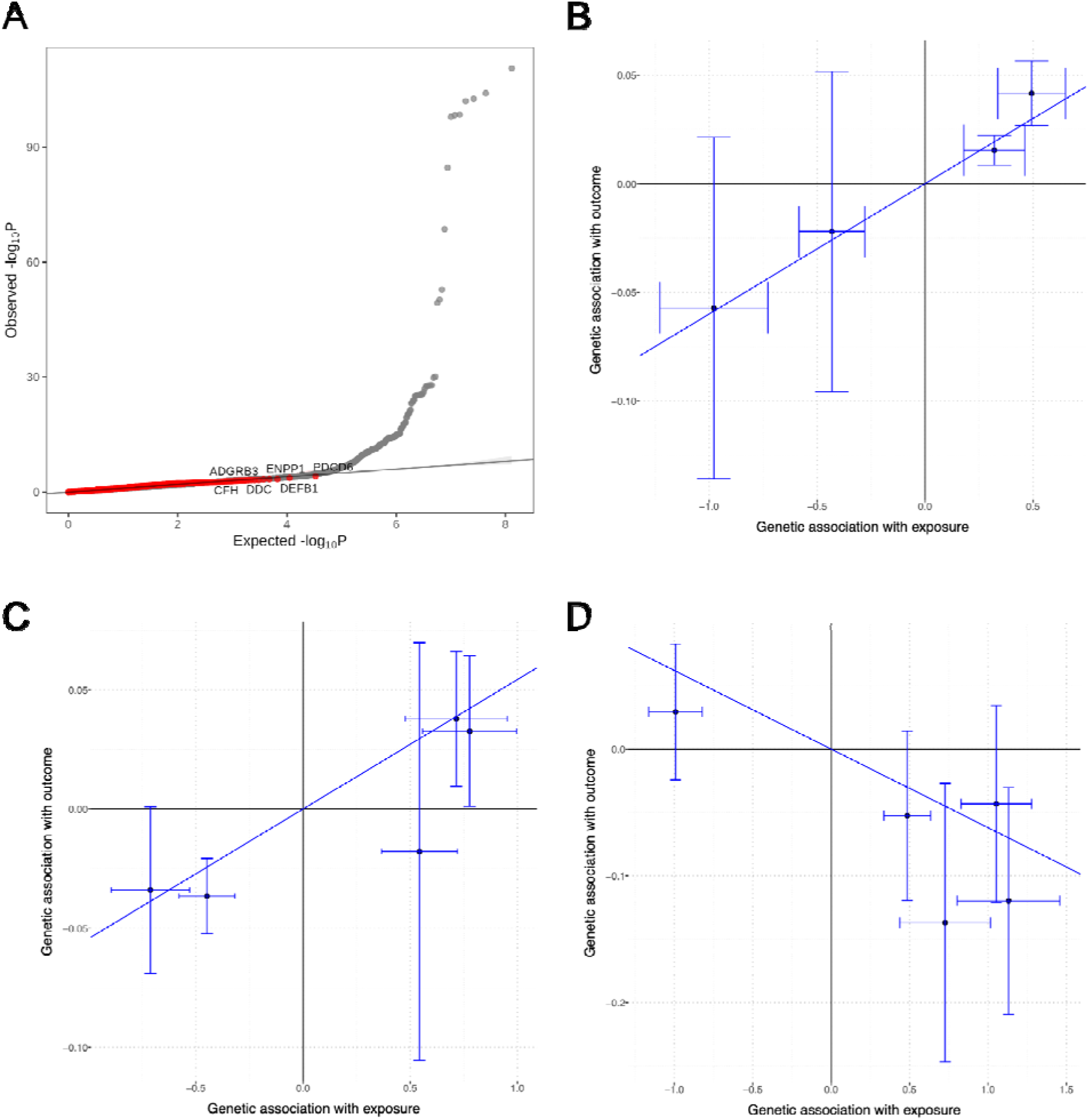
Mendelian randomization (MR) nominates candidate regulators of fetal hemolgobin (HbF) production in SCD patients. (**A**) Quantile-quantile (QQ) plot of genome-wide association results for HbF levels in 3,141 SCD patients. Red points indicate variants that reach genome-wide significance in our GEN-MOD pQTL analysis. The inflation of the test statistics is due to genetic variants at three known HbF-associated loci (*BCL11A*, *HBS1L-MYB*, *HBB*). MR results for (**B**) PTX3, (**C**) ENPP5 and (**D**) HbZ. Each point corresponds to a pQTL variant effect size for the exposure (*x*-axis, protein) and outcome (*y*-axis, HbF) with their corresponding standard errors (horizontal and vertical whiskers).

**Table 2.** Prioritization of plasma proteins implicated in the regulation of fetal hemoglobin (HbF) levels by Mendelian randomization (MR). To be prioritized, proteins need to have at least four conditionally independent pQTL (<500-kb from transcription start site), no heterogeneity (inverse variance weighted [IVW] heterogeneity QQ P-value>0.05), a corrected IVW P-value<0.05 and a nominal P-value<0.05 in the simple median MR sensitivity test. In bold, we highlight the five proteins that are associated with increased HbF levels. FDR, false discovery rate.

| Protein | Number of pQTL | IVW het. (QQ) P-value | IVW estimate | IVW FDR | Simple median P-value | Protein description |
| --- | --- | --- | --- | --- | --- | --- |
| CFHR2 | 8 | 0.51 | -0.0831 | $2.0 \times 10^{-12}$ | $1.1 \times 10^{-5}$ | Complement factor H related 2, complement cascade and innate immunity |
| FRZB | 4 | 0.76 | -0.0419 | $4.1 \times 10^{-10}$ | 0.012 | Frizzled related protein, Wnt pathway agonist |
| **PTX3** | 4 | 0.29 | 0.0600 | $9.1 \times 10^{-9}$ | 0.0076 | Pentraxin 3, marker of inflammation, elevated in SCD patients |
| VSTM1 | 7 | 0.18 | -0.0609 | $2.9 \times 10^{-8}$ | 0.020 | V-set and transmembrane domain containing 1 |
| **ENPP5** | 6 | 0.52 | 0.0544 | $3.8 \times 10^{-8}$ | $3.2 \times 10^{-4}$ | Ectonucleotide pyrophosphatase/phosphodiesterase family member 5, nucleotide metabolism |
| **ZP3** | 5 | 0.69 | 0.0395 | $9.6 \times 10^{-6}$ | 0.015 | Zona pellucida glycoprotein 3, sperm-oocyte binding |
| ALPP | 4 | 0.96 | -0.0551 | $5.5 \times 10^{-5}$ | 0.0020 | Placental alkaline phosphatase |
| GAL | 4 | 0.45 | -0.0410 | $8.3 \times 10^{-4}$ | 0.010 | Galanin, neuroendocrine peptide |
| FCRL2 | 7 | 0.19 | -0.0232 | 0.0032 | 0.021 | B-cell receptor-like protein |
| **LBP** | 4 | 0.99 | 0.0553 | 0.015 | 0.042 | Lipopolysaccharide binding protein, innate immune response |
| HBZ | 5 | 0.20 | -0.0621 | 0.023 | 0.0016 | Hemoglobin subunit zeta, part of embryonic hemoglobin |
| **NAAA** | 5 | 0.14 | 0.0510 | 0.026 | 0.023 | N-acyl ethanolamine acid amidase |
| CLEC5A | 4 | 0.37 | -0.0463 | 0.027 | 0.048 | C-type lectin domain containing 5A |
| PCDH9 | 4 | 0.13 | -0.0615 | 0.027 | 0.045 | Protocadherin 9, cell adhesion in neural tissues in the presence of calcium |

Intriguingly, we also found that genetically controlled levels of HBZ, the zeta subunit of embryonic hemoglobin, were negatively associated with HbF levels in our MR analyses (**Fig. 3D**). During human pre-natal development, embryonic hemoglobin is the first hemoglobin to be produced and is succeeded by a switch to HbF production around 10-12 weeks. The regulation of embryonic hemoglobin has not been thoroughly studied, and it is unknown whether increasing its production would result in a reduced synthesis of HbF, especially in the context of stress erythropoiesis observed in SCD (Liu et al. 2024).

## DISCUSSION

While genetically homogeneous - caused by only a small number of mutations in the β-globin gene - SCD is an extremely heterogeneous disorder that can affect multiple organs at variable severity levels (Kato et al. 2018). Understanding and modulating the genetic, environmental and socioeconomic factors that influence severity in SCD could help develop precision prevention and medicine strategies for these patients. In particular, targets that are supported by human genetic evidence are more likely to yield effective drugs (Nelson et al. 2015; King et al. 2019; Minikel and Nelson 2025; Minikel et al. 2020; Trajanoska et al. 2023), as shown by the approval of the CRISPR-based gene therapy that disrupts *BCL11A*, a negative regulator of HbF discovered by GWAS (Menzel et al. 2007; Uda et al. 2008; Lettre et al. 2008). Because proteins represent the main class of drug targets, we reasoned that a proteogenomic experiment could support the development of new therapeutic strategies in SCD.

We measured 5,411 plasma proteins in 343 SCD patients and identified 560 pQTL at genome-wide significance. Although the sample size of our experiment was at least one order of magnitude smaller than the latest pQTL studies carried out in non-SCD populations, we found 58 (10%) novel pQTL that implicated 54 unique proteins. The main reason to explain these novel findings is that we used the latest Olink HT platform, which includes ∼2,000 proteins that were not measured in previous iterations. Given that most pQTL have similar effect sizes across populations and conditions (**Fig. 2**), our results provide new MR instruments for 54 plasma proteins that can now be tested for causality in various diseases.

We found a few examples where the pQTL effect sizes in SCD patients are different than in the JHS (**Table 1**). This is a critical observation in the context of MR assumptions. To illustrate this point, consider the *trans*-pQTL signal upstream of the *HP* gene. Haptoglobin is a key molecule that scavenges cell-free hemoglobin released in the bloodstream due to hemolysis in SCD. Because high cell-free hemoglobin increases oxidative stress and reduces nitric oxide bioavailability (Kato et al. 2017), a reasonable hypothesis would be that a drug that increases haptoglobin concentration would reduce SCD severity. A “standard” strategy would be to use as MR instruments pQTL discovered in large and mostly non-diseased cohorts (e.g. UK Biobank, JHS), and test them for causal effects on SCD complications. However, our results suggest that this strategy would not work for the *HP trans*-pQTL because SCD seems to mask its effect through epistatic interactions. We have shown a similar effect previously for the association between the Duffy null variant and its association with neutrophil counts in SCD (Pincez et al. 2023). To obtain valid MR results, it is critical to confirm the strength of the instruments for the exposures (e.g. pQTL) in the outcome populations (e.g. SCD or any other human complex diseases).

Building on our pQTL discoveries in SCD patients, we used MR to identify plasma proteins that can potentially increase HbF production (**Table 2**). Of the five prioritized proteins, ENPP5 is arguably the most promising candidate (**Fig. 3B**). ENPP5 belongs to the ecto-nucleotide pyrophosphatase/phosphodiesterase (ENPP) family, which includes seven members (ENPP1–7) in the human genome. ENPP enzymes have been implicated in bone mineralization, fibrotic diseases, and immune responses, although the specific functions of ENPP5 remain unknown (Borza et al. 2022). ENPP5 (but also ENPP1, ENPP2 and ENPP4) hydrolyzes extracellular nucleotides (Gorelik et al. 2017). This is interesting because hydroxyurea, the main drug used in SCD to induce HbF production, is a ribonucleotide reductase inhibitor that stresses erythropoiesis by depleting the dNTP pool necessary for DNA replication (Orah S. Platt 2008). Additional experiments should try to confirm our MR result and establish if ENPP5 modulates HbF production by regulating the nucleotide pool.

In this study, we present a genetic characterization of the plasma proteome in SCD patients. We identified novel pQTL that can become important instruments in causal inference analyses (e.g. MR) carried out in both SCD and non-SCD populations. We also highlight five proteins as potential targets to modulate HbF levels, the main modifier of SCD severity. These results will require functional validation in experimental cellular and animal models. Together, our findings can support the development of drug development pipelines in SCD, a particularly important goal given the lack of approved molecules, recent increase in mortality rate (Abraha et al. 2025), and the expected increase in the number of SCD patients in the coming decades (Piel et al. 2014).

## METHODS

### Ethics

We collected data according to the Helsinki declaration and the study was approved by the Montreal Heart Institute ethics committee, Project #2009-106 (09-1137).

### Quantification of plasma protein levels in GEN-MOD

The levels of 5,416 proteins were measured with Olink Explore 5K in 343 participants with the SCD genotype HbSS that also have genome-wide genotyping data available. Plasma protein levels were quantified, quality checked and filtered as described in Chignon et al. 2025. To construct gaussian phenotypes for GWAS, we performed inverse normal transformation on plate control normalized NPX values for each protein.

### Mapping of protein quantitative loci (pQTL)

The GEN-MOD genome-wide genotyping data, its quality-control steps and imputation using TOPMed haplotypes has been described elsewhere (Ilboudo et al. 2025). To map pQTL, we ran `plink2 --linear` (Chang et al. 2015) on each protein level and included a total of 22 covariates: sex, age and the first 20 principal components to account for population structure. pQTL pairs were divided into *cis* and *trans*, with *cis* pairs featuring the variants within 1 Mbp of the gene encoding the associated protein, while all other pairs being labeled as *trans*. Significant *cis*-pQTL were thresholded at P-value<5×10^−8^ and *trans*-pQTL at P-value<7.7×10^−11^ in accordance with Tahir et al. 2024.

### Conditional analyses of pQTL

To identify independent sentinel variants for each protein, we performed conditional analysis with individual-level genotypes and phenotypes on all significant hits from plink2 (see https://github.com/cgroza/cond/blob/main/cond.py). To achieve this, we rank all significant hits for a protein from the smallest to the largest P-value. Then conditioning on the most significant variant, we rerun linear regression for the remaining variants, obtaining a new set of conditional P-values. Variants that lose significance upon conditioning are considered dependent on the conditioning variant. The conditioning variant is considered an independent variant for this protein. Then, the conditioning variant and its dependent variants are removed from the ranking. If any variants remain significant after conditioning at *cis/trans* genome-wide P-value thresholds, we repeat the above process recursively until no independent variants remain. We include the original 22 covariates in the conditional analyses.

### Replication of previous findings

We counted the number of replicated JHS pQTL (Tahir et al. 2024) in GEN-MOD at genome-wide significance thresholds (*cis* and *trans*) and at a relaxed P-value threshold of P-value<5.4×10^−5^. We determined the relaxed threshold through a Bonferroni correction for the number of significant variant-protein pairs in JHS that we matched with GEN-MOD (N=926).

We retrieved gnomAD (Karczewski et al. 2020) African and non-Finish European (NFE) allele frequencies for each pQTL using https://github.com/cgroza/query-gnomad.

To find variant-protein pairs with heterogeneous effect sizes between JHS and GEN-MOD, we utilized the Wald test for the difference between two independent estimates:

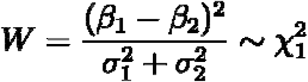

We rejected the null hypothesis if P-value(W)<1×10^−6^.

To find novel pQTLs, we matched our findings to the JHS and UK Biobank pQTL (SynID: syn51365308) tables by SNP identity and LD. Any pQTL for which the assay was not tested in the JHS or UK Biobank, or the SNP was not tested and not in LD (*r*^2^<0.2) with a reported JHS or UK Biobank pQTL variant was considered novel. We obtained UK Biobank pQTL from Synapse (Arnold et al. 2023). We obtained JHS pQTL from supplementary tables in Tahir et al. 2024.

### Genetic associations between GEN-MOD pQTL and SCD-related complications

We tested the association between the 560 pQTL discovered in GEN-MOD and five SCD-related complications: stroke, priapism, leg ulcers, avascular necrosis and cholecystectomy. For these lookups, we combined association results from GEN-MOD and the Cooperative Study of Sickle Cell Disease (CSSCD), limiting analyses to participants 12 years or older at baseline, except for stroke where we took all participants (**Table S1**). We performed association testing under an additive genetic model in SAIGE, correcting for age, sex and the first 10 genetic principal components (Zhou et al. 2018). We meta-analyzed association results with metal (Willer et al. 2010).

### Phenome-wide association study (PheWAS) of GEN-MOD pQTL

We performed PheWAS on the independent pQTL variants against 159 laboratory results from the Million Veteran Program (MVP)(Gaziano et al. 2016). To choose significant PheWAS hits, we thresholded the MVP at P-value<5.77×10^−7^, calculated through a Bonferroni correction for the number of MVP labs (n=159) multiplied by the number of independent GEN-MOD pQTL variants (545, autosomes only since the X-chromosome was not tested in MVP). Similarly, we checked the independent pQTL variants against a meta-analysis for HbF levels involving several SCD cohorts but not GEN-MOD (Ilboudo et al. 2025).

### Mendelian randomization

To select instrumental variables for MR analyses, we ran conditional analysis for *cis* pQTLs, keeping all independent SNP-protein pairs where P-value<1×10^−3^ and the distance to the transcription start site was less than 500-kb. For downstream analyses, we only kept proteins with at least 4 instruments. MR analyses were performed using the MendelianRandomization R package. Causal inference was evaluated using three independent methods: the inverse-variance weighted (IVW), the simple median (SM) — less sensitive than IVW to the distribution of the IVs — and Cochran’s Q test to assess heterogeneity and potential horizontal pleiotropy. A Benjamini–Hochberg false discovery rate (FDR) correction was applied to IVW P-values. Proteins were prioritized as putatively causal if they met the following criteria: IVW FDR<0.05, SM P<0.05, and Cochran’s Q test P>0.05.

## ACKNOWLEDGMENTS

We thank all GEN-MOD participants who contributed data to this study. We also thank Pouria Jandaghi and Daniel Auld from the McGill Genome Center. We thank Wenmin Zhang for statistical support. We acknowledge the support of the Digital Research Alliance of Canada (https://www.alliancecan.ca/en).

## AUTHOR CONTRIBUTIONS

Conceived and designed the analyses: C.G. and G.L.; Collected the data: A.C. and P.B.; Contributed data: P.B.; Performed analyses: C.G., A.C., K.S.L., V.B. and G.L.; Secured funding and supervised the work: G.L.; Wrote the manuscript: C.G., A.C. and G.L., with contributions from all authors.

## DATA AVAILABILITY

The GEN-MOD proteomic data has not been deposited in a public repository because the data is not public but is available from the corresponding author on request. Scripts to analyze the data and draw figures are available on Zenodo (Groza 2026).

